# Endocrine and well-being profiles shape psychosexual health and psychophysiological symptoms after menopause

**DOI:** 10.64898/2026.10.06.26364576

**Authors:** Anna F. Denninger, Jule Sippel, Sara Beck, Amelie Decker, Amelie Walther, Pia Büttner, Anne Starke, Inger Sundstöm-Poromaa, Melanie Henes, Birgit Derntl, Lydia Kogler

## Abstract

Menopause is a major neuroendocrine transition characterised by substantial changes in endocrine milieu, physiological, psychological, and psychosexual health. Although menopausal symptoms are common, their range and severity vary considerably between individuals, suggesting that menopause is a multifaceted process. Endocrine markers (luteinising, follicle-stimulating & anti-Müllerian hormone), together with perceived well-being, may contribute to this variability. Thus, the aim was to capture the complex impact of endocrine and well-being markers on psychosexual health and menopausal symptoms. Therefore, we first applied principal component analyses to identify individual endocrine and well-being profiles, to then assess their association with psychosexual health and psychophysiological symptoms in pre- and post-menopausal females. Post-compared to pre-menopausal females report reduced psychosexual health and greater urogenital and psychological symptom burden. Endocrine and well-being profiles interact with reproductive status: Post-menopausal females with lower subjective well-being and a stronger menopause-like endocrine milieu report decreased overall psychosexual health and more sexual related pain. Additionally, endocrine milieu interacted with reproductive status on urogenital symptoms: Post-menopausal females with a stronger menopause-like endocrine milieu report less urogenital symptoms.

---

Together, these findings suggest that endocrine factors and subjective well-being jointly influence psychosexual health and psychophysiological symptoms after menopause. No association is seen before menopause. Therefore, females after menopause need to be considered separately in research and clinic. Multivariate analyses capture patterns that are not identified using individual biomarkers and conventional univariate approaches. Our findings highlight the value of integrative multivariate approaches for improving the assessment of menopausal health and characterising individual variability across the menopausal transition. Trial No.: NCT06204016

Keywords: Menopause, menopause symptoms, female sexual health, endocrine markers, well-being, multivariate analysis, mental health.

## Background

Menopause represents a key neuroendocrine transition phase marking the end of the reproductive period and is often accompanied by a range of conditions impacting quality of life and well-being (e.g., mood disturbances). These include somatic complaints like vasomotor (e.g., hot flashes) and genitourinary symptoms (e.g., vaginal dryness)(1). However, the severity and manifestation of these symptoms vary considerably between individuals (2). Research on menopause has largely focused on vasomotor symptoms and impairments in sexual health (e.g., vaginal dryness, reduced libido); both significantly impairing quality of life (3–5). A meta-analysis reports a twofold increase in the prevalence of symptoms such as vaginal dryness in post-compared to pre-menopausal females (6), and the prevalence of these symptoms varies with reproductive status (1).

A hallmark of menopause is the large-scale reorganisation of endocrine signalling. As ovarian reserve declines, levels of oestradiol and progesterone are markedly reduced, while gonadotropins such as follicle-stimulating hormone (FSH) and luteinizing hormone (LH) become elevated (7,8). In parallel, anti-Müllerian hormone (AMH), a marker of ovarian reserve, becomes nearly undetectable. These menopause-associated hormones seem to have distinct impacts on psychosexual health and psychophysiological complaints. In post-menopausal females, orgasm frequency has been positively associated with LH levels. In addition, higher sexual desire has been associated with better mental health, while sexual arousal, orgasm, and satisfaction with sexual life are associated with lower pain levels during post-menopause (9). In pre-menopausal females, FSH levels were positively associated with orgasm frequency, whereas LH levels were negatively correlated with pain intensity (10). Next to psychosexual health, somatic menopausal symptoms are impacted by endocrine shifts during perimenopause. Higher AMH levels have been associated with a reduced likelihood of hot flushes, whereas FSH levels show the opposite relationship (11). Moreover, lower AMH levels in the mid-30s have been associated with increased odds of vaginal dryness later in life (12). Longitudinal population-based studies following females from pre- to post-menopause over 11–14 years indicate that increases in FSH levels are associated with greater depressive symptom severity (13,14). Also depressed pre-menopausal females exhibit higher FSH levels and more severe psychophysiological symptoms, particularly during the follicular phase of the menstrual cycle (15). However, a 10-year longitudinal study found no association between FSH levels and major depressive symptoms during the menopausal transition (16). Nevertheless, it seems that LH, FSH and AMH impact the appearance of psychological and vegetative menopausal symptoms. These menopausal changes are unlikely to be driven by a single hormone alone but rather by a complex endocrine interaction. Combined biomarker profiles would be necessary to better characterize symptom variability and underlying neuroendocrine changes in individuals.

In addition to these endocrine alterations, well-being plays an important role in shaping menopausal experiences and psychosexual health throughout the reproductive life span. Well-being decreases with increasing depressive symptom severity and stress (17). Although some studies report greater depressive symptom severity in postmenopausal than in premenopausal women (18), other cross-sectional studies have found no significant differences in the prevalence of depressed mood across hormonal stages (19–21). Beyond these differences across the menopausal transition, depression is also associated with poorer psychosexual health (22,23), with greater depressive symptom severity correlating with poorer psychosexual well-being and increased sexual pain (24). These associations may be particularly pronounced during perimenopause and post-menopause, when higher depressive symptom levels coincide with poorer psychosexual health compared with pre-menopause (25). Similarly, while perceived stress has been shown to exacerbate menopausal symptoms (26), evidence for a direct association between menopausal status and stress levels remains inconclusive: post-compared to pre-menopausal females report comparable levels of well-being including perceived stress, anxiety, and depression (27). Nevertheless, a substantial body of research indicates that higher perceived stress is associated with greater menopausal symptom burden, including poorer well-being, increased psychological and more frequent and persistent vasomotor symptoms (3,26,28–30). Additionally, in females in early midlife (44–55 years), increased urogenital, somatic, and psychological symptoms are associated with elevated anxiety, underscoring the close interrelationship between menopausal symptoms and well-being (31).

Both endocrine and well-being factors contribute to psychophysiological symptoms and psychosexual health before and after menopause. However, previous research has often examined individual hormones or well-being domains in isolation. Such univariate approaches fail to capture the complex, reciprocal interactions within the endocrine system and the multidimensional nature of subjective well-being, potentially limiting our understanding of the diverse symptoms and symptom burden associated with menopause.

Consequently, clinical practice and research increasingly recognises the importance of combining biological (e.g. endocrine, inflammatory & metabolic markers) and well-being (e.g. mental health, stress, emotion regulation) markers. Such approaches have improved risk prediction in cardiovascular disease, enhanced diagnostic accuracy in ovarian cancer and depression subtype characterisation, demonstrating the value of combining biological signals rather than examining markers in isolation (32–34). This integrative approach is especially relevant in female health, where changing endocrine and psychological processes are additionally shaped by dynamic interactions (35). Across the menstrual cycle and menopausal transition, rapidly fluctuating hormone levels influence multiple interconnected systems, making it unlikely that single markers alone can adequately explain symptom variability. Multivariate analyses are therefore necessary to disentangle these complex relationships and identify patterns that are based on multiple variables.

Thus, we want to assess the impact of the endocrine and well-being variables on psychophysiological symptoms and psychosexual health. We used a two-step approach by applying principal component analyses to reduce multivariate complexity, followed by linear mixed models assessing the impact of endocrine and well-being profiles on menopausal symptoms and psychosexual health. We hypothesise that post-compared to pre-menopausal females would report greater menopause-related physiological and psychosexual symptom burden, and that endocrine and well-being profiles would be associated with menopausal symptom and sexual health dependent on reproductive status (pre-vs. post-menopausal), whilst controlling for age. Exploratorily, we investigated whether these profiles and reproductive status also jointly interact on menopausal symptoms and psychosexual health, as the complex interplay between endocrine changes and well-being remains insufficiently understood.

## Methods

### Sample

A total of 80 biological females (sex assigned at birth) participated in the study. All participants required to be aged between 18-70 years, non-smokers, and fluent in German. Pre-menopausal females (> 18 years) were naturally cycling with regular menstrual cycles (25–35 days,), while post-menopausal females reported at least ≥ 12 months of amenorrhea (7).

Exclusion criteria comprised current or past endocrine, cardiovascular, neurological, metabolic (e.g., diabetes, severe hypertension) or mental disorders (*Structured Clinical Interview for DSM-5*; (36)), current pregnancy and breastfeeding within the past year, recent childbirth, and hormonal therapy as well as oral contraceptive use within the last three months.

The study was embedded in a larger preregistered study (registration number: NCT06204016) and approved by the ethics committee of the University of Tübingen (No.: 426/2023B02). All participants provided written informed consent prior to participation.

### Design

After screening for eligibility, participants provided sociodemographic information, questionnaires assessing psychosexual health (FSFI; (37)), menopausal symptoms (MRS; (38)), depressive symptoms (BDI; (39)), quality of life (WHO-5; (40)), state and trait anxiety (STAI; (41)), perceived stress (PSS-10; (42)), amongst others variable that are relevant for the broader study (NCT06204016). Additionally, blood samples were collected. For pre-menopausal females, blood samples were drawn during the early follicular phase (days 2–5 following menstruation onset) to control for hormonal fluctuations.

Subjective data were collected and stored using REDCap electronic data capture tools hosted at meDIC / University Hospital Tuebingen (43).

### Hormone analyses

Serum hormone levels of LH, FSH and AMH were analysed by the Zentrallabor Tübingen applying chemiluminescence immunoassay (CLIA) with sensitivity ranges as follows: LH (.07 IU/l), FSH (.3 IU/l) and AMH (.31 pmol/l). Hormone concentrations below the assay detection limit (AMH: .01 or .03 pmol/l) were replaced with the respective detection-limit value for further analysis.

### Statistical analyses

All data analysis was performed with MATLAB 2024a. Group comparisons were performed using Welch-testing. If normality was not met, Mann-Whitney U-tests were applied. Chi² test was applied for comparisons of categorical data.

### Principle component analyses (PCA)

To reduce the complexity of impacting variables, we applied principal component analyses for (a) the endocrine milieu and (b) for subjective well-being. Therefore, an initial set of variables were tested for multi-collinearity (Pearson correlation). Then Bartlett’s test of sphericity and the Kaiser–Meyer–Olkin (KMO) test were performed to assess the suitability of the data for PCA, and to determine whether individual variables can be retained (KMO > 0.57). This procedure resulted in the following variables that can be included in the PCA: (a) for the endocrine milieu: LH, FSH and AMH, and (b) for subjective well-being: trait anxiety (STAI), depressive symptoms (BDI-2), perceived stress sub-scores self-efficacy and hopelessness (PSS-10), and quality of life (WHO-5). All variables, except for binary group allocation (reproductive status = pre-vs. post-menopausal), were z-transformed prior to analysis. Principal components were retained based on eigenvalues greater than one and inspection of the scree plot. To further test the effect of group (pre-vs. post-menopause) and individual endocrine (endoPCs) and well-being (wellPCs) profiles on menopausal symptoms and psychosexual health we fitted general linear-mixed models. Therefore, PC-scores of each profile (endocrine and well-being) were added in the model as predictor as previously done (e.g., (44)):

### Symptoms (psychosexual health or menopausal) ∼ groups * endoPC_scores *wellPC_scores + age

All analyses were performed after outlier removal (3 > residuals < 3). False discovery rate (FDR) correction (Benjamini–Hochberg procedure) was applied, correcting in total across 4 subscales of the menopausal rating scale (MRS) and 7 subscales of the female sexual functioning index (FSFI). In case of significant 3-way interactions, the post-hoc tests were examined with simple slope analyses: Therefore, we separated data in groups based on the standard deviation (SD) of the endocrine or well-being profile (low scores: -1SD, mean scores: SD=0, and high scores: +1SD) which is indicating either a mean profile (SD=0), a profile below the mean (−1SD) or above the mean (+SD). This was done separately for pre-and post-menopausal females while controlling for age.

## Results

### Sample Description

In total 45 pre- and 33 post-menopausal females were included. Two pre-menopausal females were excluded due to the possibility of being peri-menopausal, based on age (>51 years). Age (*p < .001*), age at menarche (*p = .022*), relationship status (*p = .001*), education (*p <.001*) and nett income (*p <.001*) were significantly higher in post-compared to pre-menopausal females. Depressive symptoms (*p = .020*), and perceived stress (*p_hopelessness_ = .032; p_self-efficacy_ =.013; p_total_ = .0185*) were higher in pre-compared to post-menopausal females (Table 1). However, groups did not differ in quality of life, state and trait anxiety (*p_quality_ = .060; p_state_ = .213; p_trait_ = .090*).

**Table 1.**
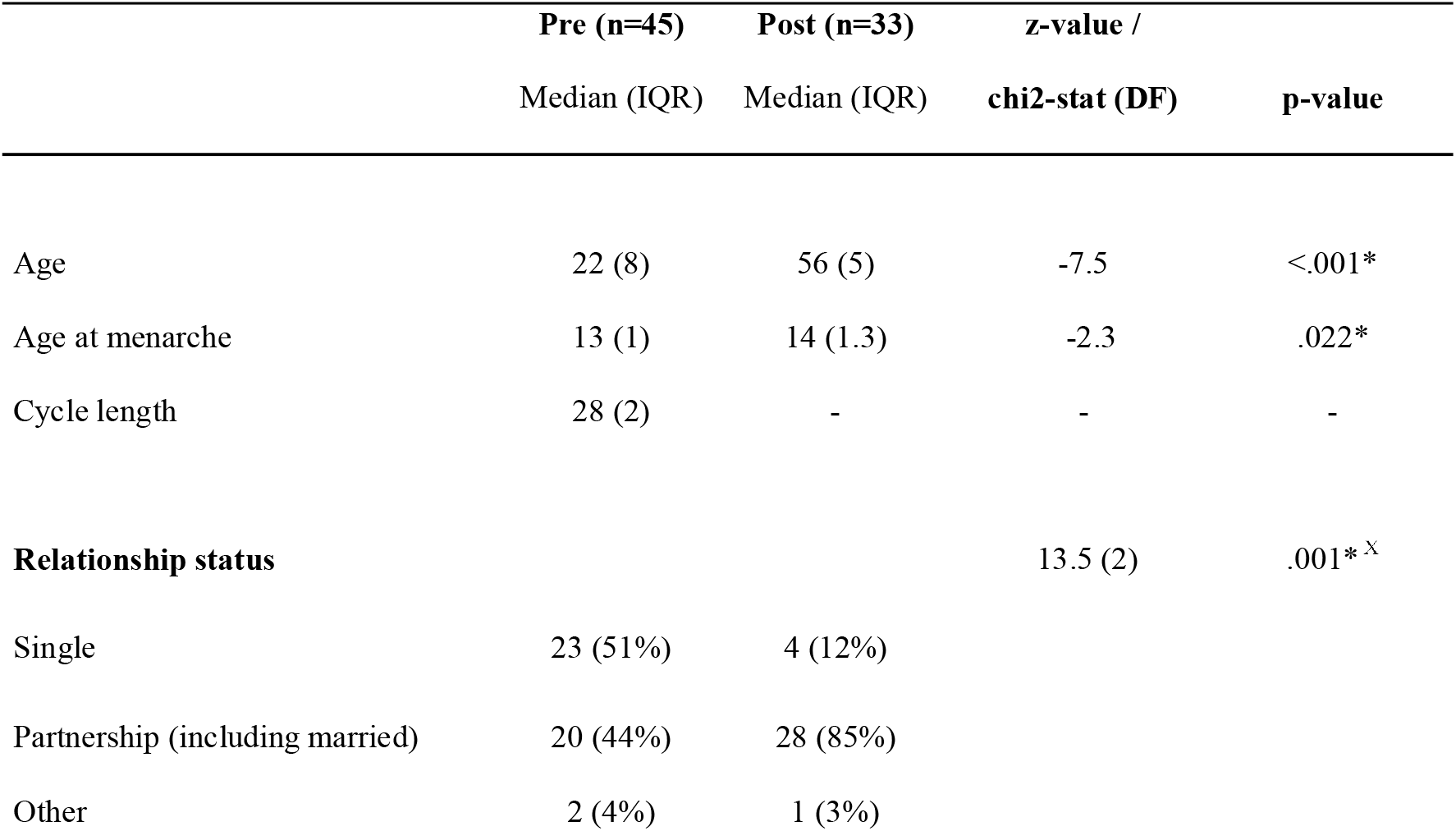
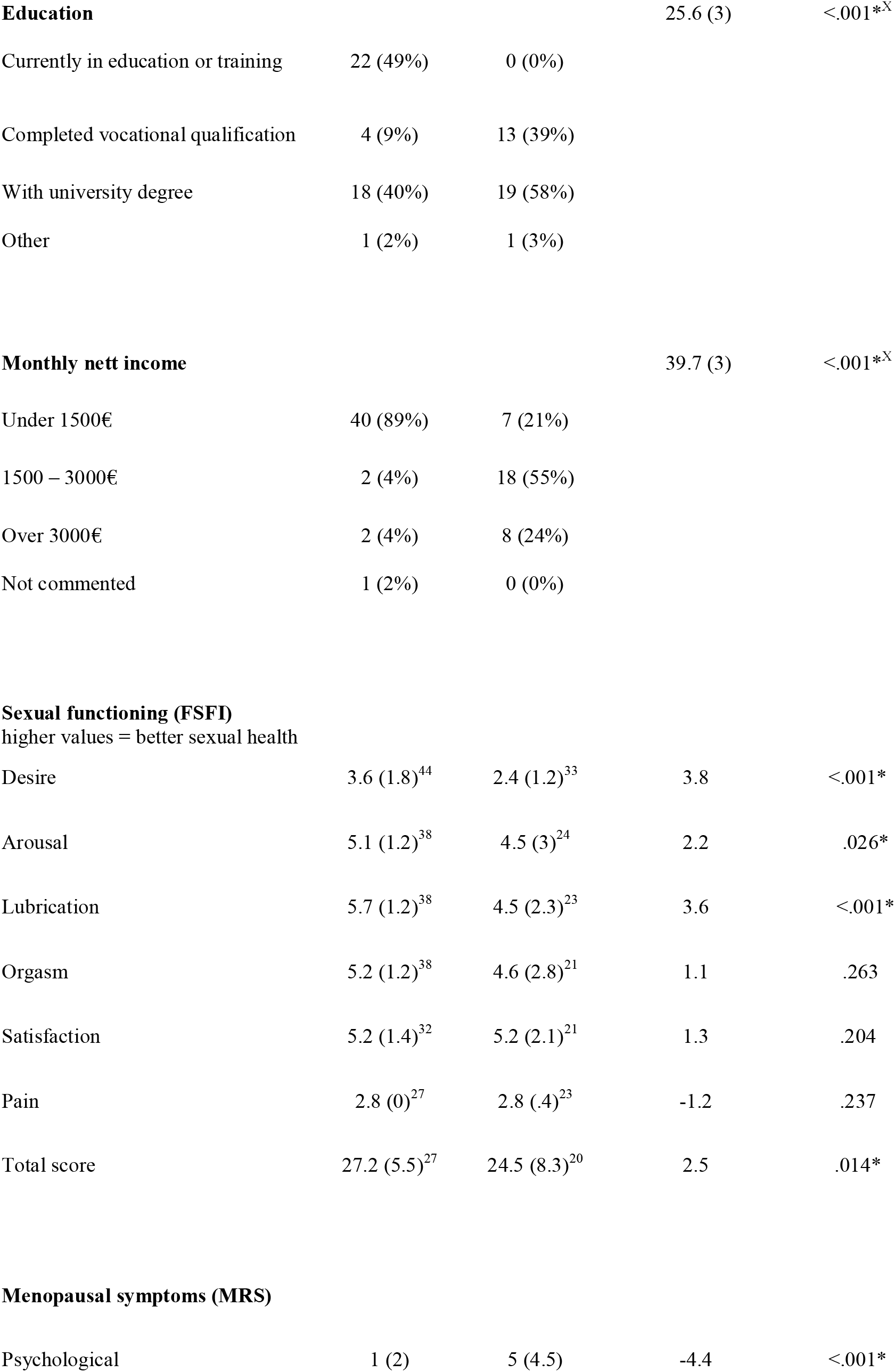
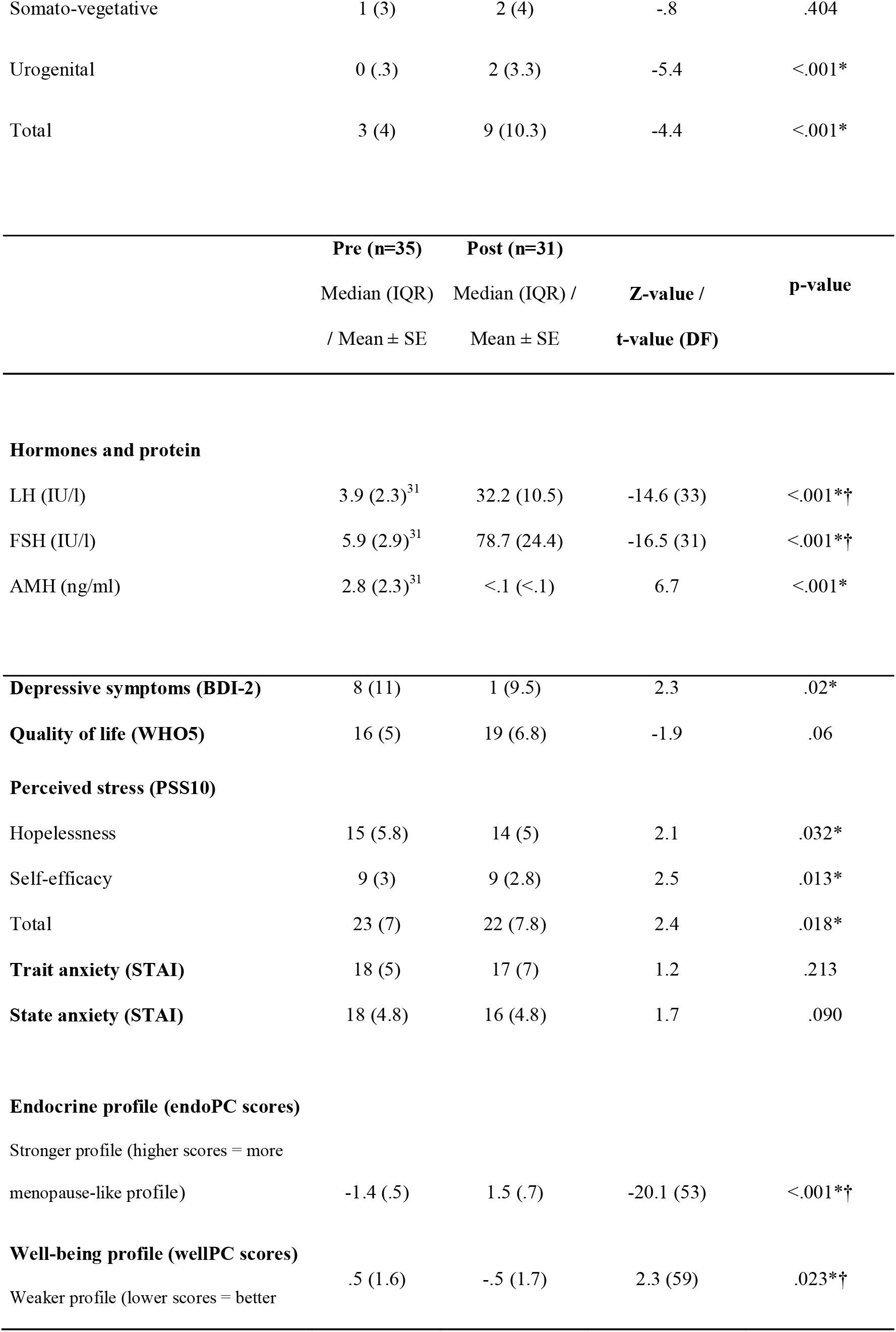
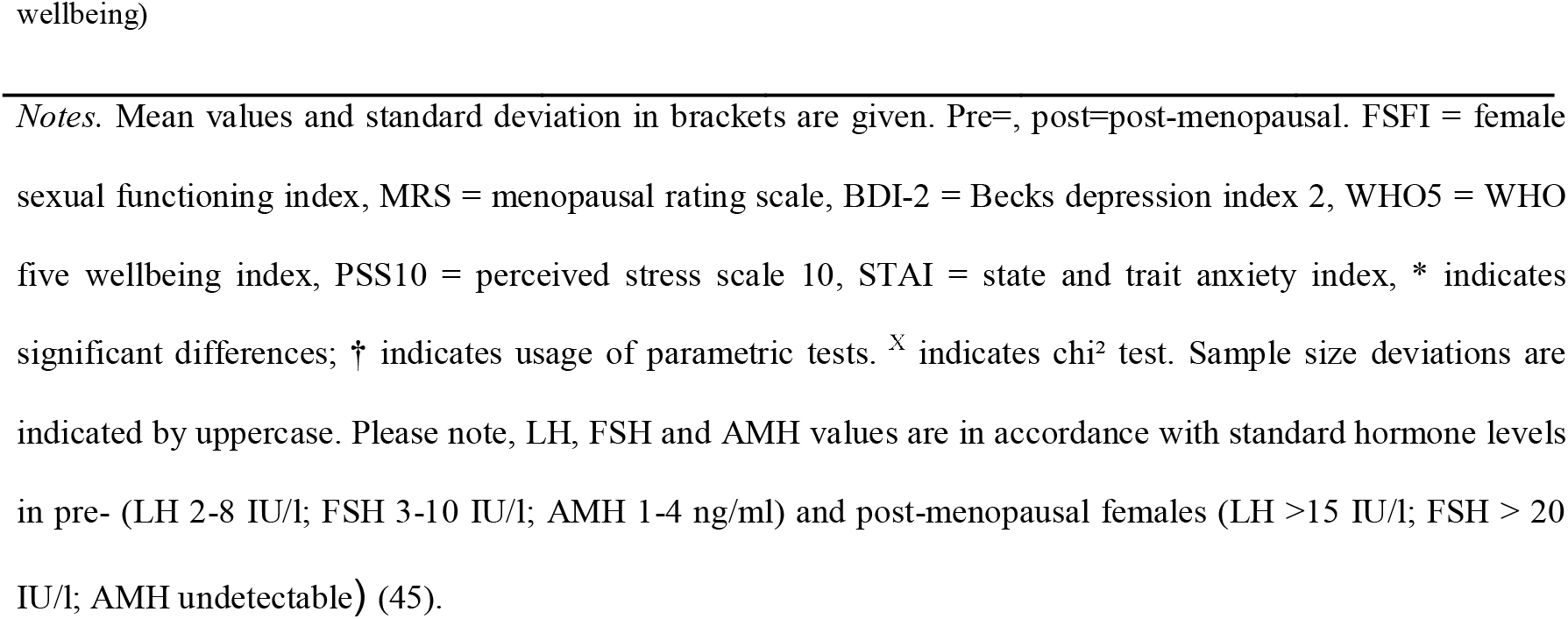
Sample description. Notes. Mean values and standard deviation in brackets are given. Pre=, post=post-menopausal. FSFI = female sexual functioning index, MRS = menopausal rating scale, BDI-2 = Becks depression index 2, WHO5 = WHO five wellbeing index, PSS10 = perceived stress scale 10, STAI = state and trait anxiety index, * indicates significant differences; † indicates usage of parametric tests. ^Χ^ indicates chi² test. Sample size deviations are indicated by uppercase. Please note, LH, FSH and AMH values are in accordance with standard hormone levels in pre-(LH 2-8 IU/l; FSH 3-10 IU/l; AMH 1-4 ng/ml) and post-menopausal females (LH >15 IU/l; FSH > 20 IU/l; AMH undetectable) (45).

Psychosexual health also differed between groups: pre-compared to post-menopausal females exhibited significantly higher scores of desire, arousal, lubrication and total score (*p_desire_ <.001; p_arausal_ = .026; p_lubrication_ >.001; p_total_ = .014*), indicating better psychosexual health. Psychological, urogenital and total menopausal symptoms were significantly more pronounced in post-than pre-menopausal females *(all p<.001*). Somato-vegetative symptoms did not differ between groups (*p = .404)* (Table 1).

### Hormone and protein data

As expected, post-compared to pre-menopausal females exhibited significantly higher concentrations of LH, and FSH, but lower levels of AMH (all *p* < .05).

#### Principal component analyses

For both endocrine and well-being profiles, the PCA identified one principal component (eigenvalue > 1): explaining 82.4% of the variance for the endocrine (endoPC), and 73% for the well-being (wellPC) component (Figure 1).

**Figure 1.**
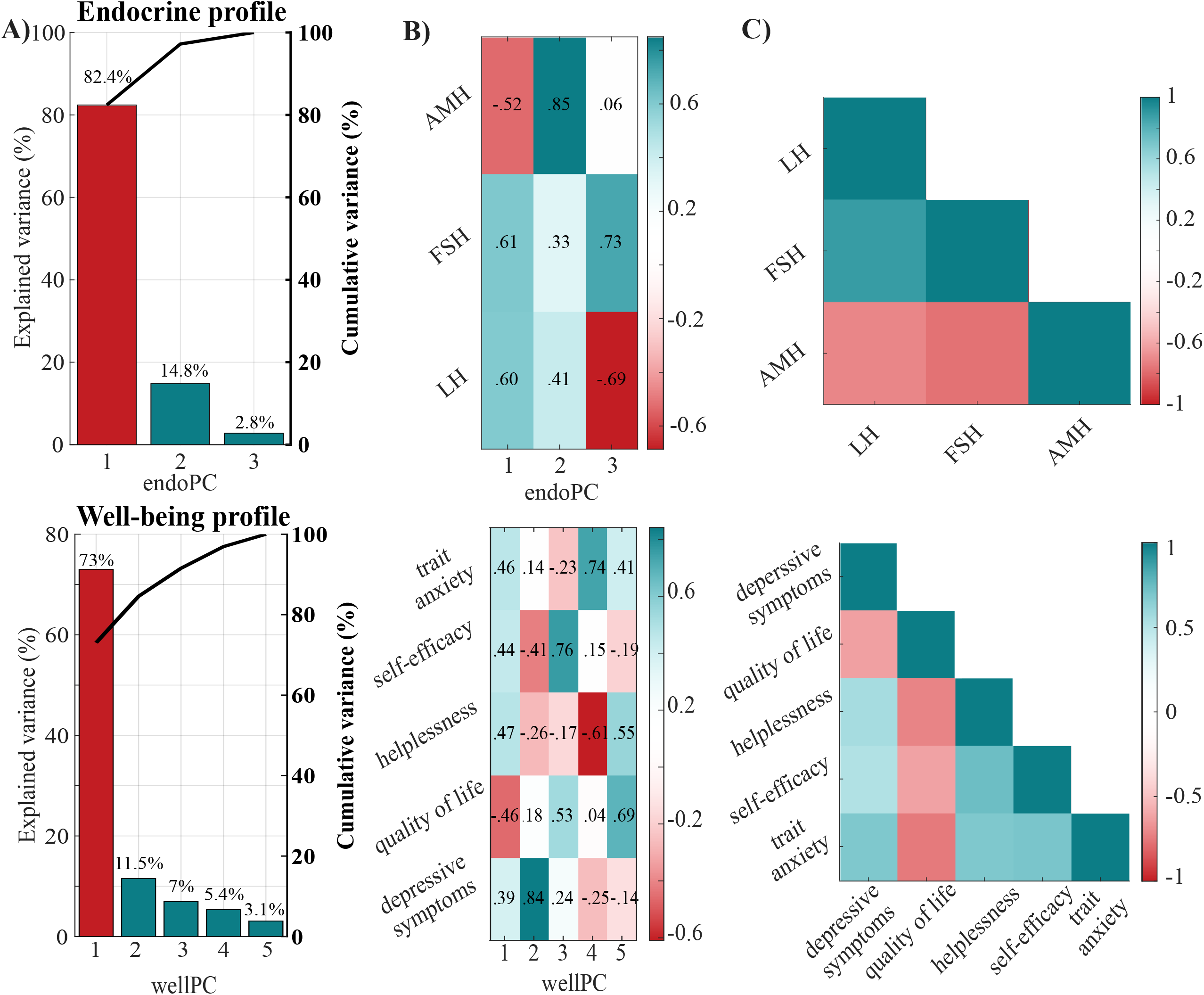
Multivariate structure of endocrine and well-being profiles. Scree plots (A) depict the proportion of variance explained by each principal component as well as the cumulative variance explained across components. Components with eigenvalues greater than 1 are highlighted in red, indicating retention under the Kaiser criterion. Principal component loadings (B) are visualized, with warm colours representing negative loadings and cool colours representing positive loadings, highlighting the direction and magnitude of variable contributions to each component. (C) A heat-map displays pairwise Pearson correlations among variables all above ≥ .5. Warm colours indicate negative correlations, while cool colours indicate positive correlations.

#### Endocrine profile

endoPC loaded positively on LH (.60) and FSH (.61), and negatively on AMH (−.52) (Figure 1). Thus, higher scores of the endoPC are indicating a post-menopausal like endocrine milieu.

#### Well-being profile

WellPC loaded positively on trait anxiety (.46), helplessness (.47), self-efficacy (.44) and depressive symptoms (.39), and negatively on quality of life (-.46) (Figure 1). Thus, lower scores of the wellPC indicate a better well-being.

Scores of endoPC (*p <.001, higher in post-menopausal females*) and wellPC (*p =.023, higher in post-menopausal females*) differed significantly between groups (Table 1).

Principle component scores of each profile, group (pre-vs. post-menopause) and their interaction were entered as predictors in linear mixed-effects models.

### Impact of endocrine and well-being profiles on psychosexual health differs between pre-and postmenopausal females

#### Sexual health

For total FSFI scores, a significant two-way interaction endoPC × wellPC (β*z = −.897, pFDR = .035*) was subsumed under a significant three-way interaction reproductive status x endoPC x wellPC (β*z = .897, pFDR = .049*). The three-way interaction indicates that the relationship between the endPC and wellPC show distinct patterns in pre- and post-menopausal females. To probe this interaction, simple slope analyses were executed separately for each group (pre- and post-menopausal females).

Simple slope analysis indicates that the association between endoPC and wellPC scores distinctly interact in post-but not premenopausal females (Figure 2). In post-menopausal females with high wellPC scores (indicating worse well-being; group: +1SD in Figure 2), endoPC scores are negatively associated with worse sexual health (group: +1SD; β *=-1.683, p = .022*). This association is not significant in post-menopausal females with mean (group: mean, 0SD; *p >.05*) and low well-being scores (indicating better well-being, -1SD*, p >.05*). Thus, in post-menopausal females with worse well-being the endocrine profile is associated with reduced psychosexual health. Among pre-menopausal females, endoPC scores are not significantly associated with psychosexual health at any level of well-being (better vs. mean vs. worse well-being; all *p >.05*) (Figure 2; supplementary table S2).

**Figure 2.**
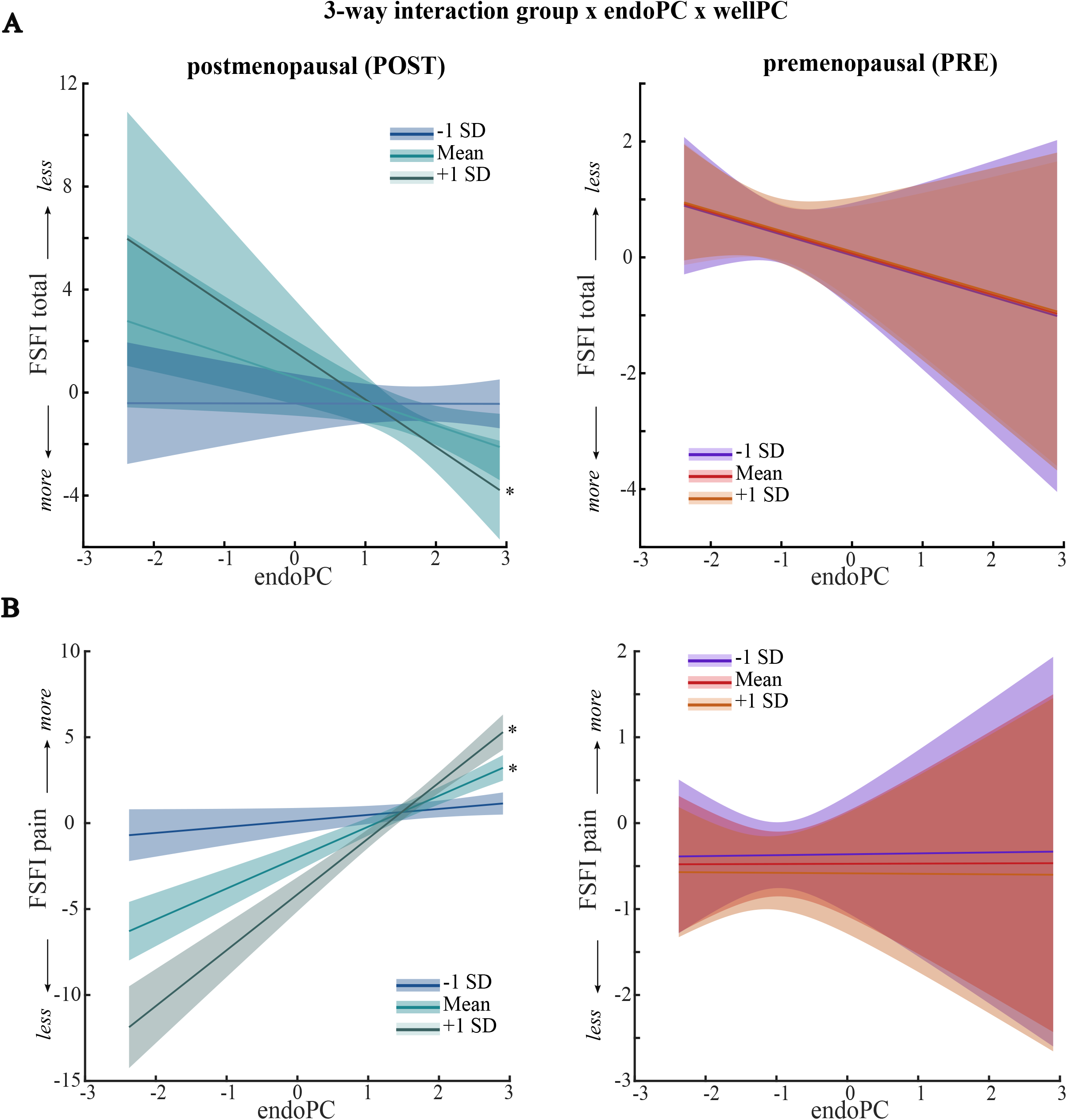
Visualisation of the simple slope analyses of the associations between endocrine and well-being profiles with female psychosexual health. For visualisation, the three-way interactions are depicted separately for females with low (group: -1SD = low profile strength = better well-being), mean (group: mean, 0SD), and high (group: +1SD = high profile strength = worse well-being) levels of wellPC scores in each group (pre-and post-menopausal) for overall FSFI (higher scores indicate worse psychosexual health) (A) and sexual related pain in the FSFI (higher scores indicate less pain) (B). Warm colours indicate pre-, and cool colours indicate post-menopausal females. Associations are presented with 95% confidence intervals. Significant associations (<.05) are indicated with an asterisk (*).

#### Pain during & after intercourse

For intercourse related pain significant main effects were observed for both endoPC (β*_z_= 1.798, pFDR < .001*) and wellPC (β*_z_ = −2.435, pFDR < .001*). Significant interactions appeared for group × endoPC (β*_z_ = −1.75, pFDR < .001*) and group × wellPC (β*_z_ = 2.305, pFDR < .001*), and group x endoPC x wellPC (β*_z_= −1.668, pFDR < .001*).

Thus, the relationship between endoPC and wellPC are dependent on reproductive status. In post-menopausal females with lower levels of well-being profiles (group: -1SD, indicating better well-being), endocrine profile scores are not significantly associated with pain scores (*p >.05*) (Figure 2). Contrarily, in post-menopausal females with mean (group: mean, 0SD) and high (group: +1SD, *n = 3,* indicating worse well-being) levels of well-being profile scores, the endocrine profile scores are significantly associated with pain during sexual intercourse scores (β *= 1.798, p >.001*; β *= 3.444, p >.001*), respectively. Thus, in post-menopausal females that report worse well-being (higher wellPC scores), a higher post-menopause endocrine profile is associated with less pain during and after sexual intercourse (Figure 2). In pre-menopausal females there is no significant association between endocrine profile, well-being profile and pain during sexual intercourse (all *p >.05*) (Figure 2; supplementary table S2).

No other main effects or interactions appeared to be significant for subscales of the FSFI after correction for multiple comparisons (supplementary table S1).

### Impact of the endocrine profile on urogenital symptoms differs between pre-and postmenopausal females

#### Urogenital symptoms

For urogenital symptoms of (MRS), significant main effects of group (β*_z_ = −1.696, pFDR = .008*) and endoPC (β*_z_ = −.846, pFDR <.001*), are subsumed under a significant interaction group x endoPC (β*_z_ = .983, pFDR = .004*). Simple slope analyses indicated that endoPC is significantly associated with urogenital symptom scores in post-(β *= -.846, p <.001*) but not in pre-menopausal females (*p > .05*) (Figure 3; Supplementary table S2). Thus, in post-menopausal females, a more menopause-like endocrine milieu (= higher endoPC scores) is associated with less urogenital symptoms. These findings suggest that the relationship between endocrine profile scores and urogenital symptoms is specific to post-menopausal females. No other main effects or interactions appeared to be significant for subscales of the MSR after correction for multiple comparisons (please see supplementary table S1 for each model)

**Figure 3.**
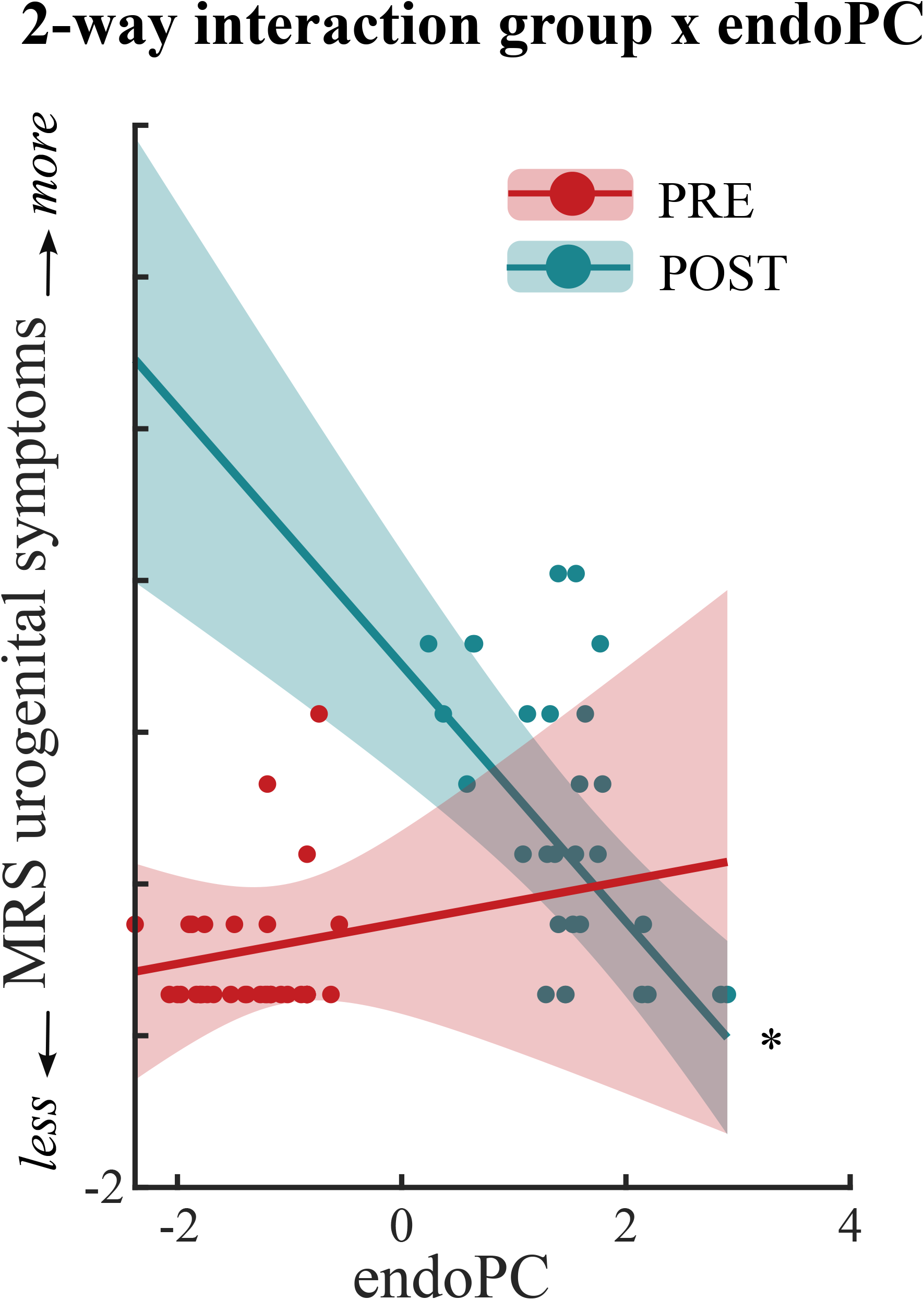
Visualisation of the simple slope analyses of the associations between endocrine profiles with urogenital symptoms. Panel C depicts the two-way interaction for urogenital symptoms between reproductive status and endoPC scores. Warm colours indicate pre-, and cool colours indicate post-menopausal females. Associations are presented with 95% confidence intervals. Significant associations (<.05) are indicated with an asterisk (*). PRE – premenopausal females; POST – postmenopausal females

### Profile models show strong and consistent explanatory performance

To assess whether the multivariate principal-component scores (endoPC and wellPC) provided additional explanatory information compared with models using individual variables (e.g., hormones or questionnaires, separately), we compared models containing single variables with models incorporating multivariate principal-component scores (see Supplementary Table S3). Overall, the endoPC × wellPC model showed consistently strong performance across all three outcomes. Please see supplementary results and table S3 for details.

Thus, the endoPC × wellPC model provided the best overall combination of explanatory performance when all three criteria (explained variance, model fit, prediction accuracy) were considered together (see supplementary Table S3 for statistics).

## Discussion

The menopause transition has substantial relevance for individual (1, 3–5) and public health (45) and will affect approximately 1.2 billion females worldwide by 2030 (46). Previous studies of menopausal health have often overlooked specific hormone concentrations, did not control specific variables (e.g. menstrual cycle phase) or assessed single endocrine and well-being variables alone. Such methodological considerations are particularly necessary considering the substantial endocrine alteration occurring during menopause and the menstrual cycle. Therefore, this study investigated differences in key subjective and endocrine characteristics in females before and after menopause and investigated their impact on psychosexual health and psychophysiological symptoms, which are key contributors to female’s health in everyday life. Firstly, our findings indicate that the association between endocrine and well-being profiles and psychosexual health differs according to reproductive status (pre-vs. post-menopause). Specifically, the endocrine and well-being profiles interact with group, indicating an association only in females after menopause. Post-menopausal females with low subjective well-being and a stronger menopause-like endocrine profile report decreased psychosexual health, but less pain related with sexual intercourse. Additionally, in post-menopausal females, urogenital symptoms are associated with the endocrine profile, independent of psychological well-being. Post-menopausal females with a stronger menopause-like endocrine profile report less urogenital symptoms. Secondly, our data indicates that multivariate principal components provide a more comprehensive representation of symptom-related variance than single individual biological and well-being markers alone. Together, these findings suggest that the interplay between endocrine and well-being differs according to the reproductive status of females. The effects of endocrine-and well-being-related factors on psychosexual health and psychophysiological symptoms are tightly coupled after menopause.

### Profile identification

In the present study, we applied multivariate analyses to characterise endocrine and psychological well-being profiles rather than relying on individual hormones or subjective measures. Principal component analysis (PCA) captures the shared variance across multiple markers, thereby identifying biologically and psychologically meaningful profiles.

### Endocrine profile

The first principal component identified for the endocrine profile reflected a post-menopausal endocrine profile, when AMH levels are low and FSH and LH levels are high (47).

### Well-being profile

For the well-being profile, again, the first principal component was identified, including subjective trait anxiety, depressive symptoms, stress coping (helplessness and self-efficacy) (all positive loadings), and quality of life (negative loading). This profile reflects a “negative well-being state” with higher levels of stress, anxiety, and depressive symptoms, together with lower quality of life.

Importantly we found that multivariate principal-component representations may provide a more comprehensive representation of variance in psychosexual and menopausal health outcomes than individual biological markers. Across all three outcomes, the endoPC × wellPC model showed consistently strong explanatory and predictive performance.

### Endocrine profile impacts psychophysiological symptoms only in post-menopausal females

#### Menopausal symptoms

Urogenital symptoms including e.g., vaginal dryness, and bladder problems were associated with endocrine profile scores in post-menopausal females. These findings suggest that, after menopause, urogenital symptoms are linked to the endocrine milieu, with less urogenital symptoms in postmenopausal females having a stronger post-menopausal-like endocrine milieu. Menopause is commonly associated with vulvovaginal atrophy and genitourinary syndrome of menopause, which are characterised by thinning of the vaginal epithelium, reduced lubrication, loss of elasticity, and increased tissue friability (48–50). The findings of the current study are supported by previous research linking reproductive hormone levels to urogenital health. For example, lower AMH concentrations in pre-menopausal females (during their mid-thirties) have been associated with increased odds of developing moderate-to-severe vaginal dryness approximately 14 years later. This suggests that markers of declining ovarian reserve may predict subsequent urogenital symptoms (12). Likewise, higher FSH concentrations have been positively associated with urogenital symptom severity in post-menopausal females (51). Associations between gonadal hormone levels and urogenital symptoms may reflect adaptation of hormone receptor availability to reproductive status (52). A more stable post-menopausal endocrine milieu may therefore require less physiological adaptation to hormonal fluctuations; however, this remains speculative and requires longitudinal investigation.

Interestingly, we found no association between the well-being profile and urogenital symptoms, which may reflect its predominant focus on vulnerability-related traits rather than protective psychological resources. Higher resilience, e.g., higher self-esteem and adaptive emotion regulation strategies, may buffer the psychological impact of menopausal complaints, whereas vulnerability-related traits such as neuroticism are associated with greater risk of depression and anxiety during the menopausal transition (27,53–58). Future research should therefore consider both psychological vulnerability and protective factors when examining individual differences in psychophysiological symptom experiences after menopause.

### Endocrine and well-being profiles impact psychosexual health in post-but not in pre-menopausal females

Psychosexual health was associated with the endocrine and well-being profile scores, again, in post-but not in pre-menopausal females. Endocrine and well-being profiles interact when impacting overall psychosexual functioning and sexual pain exclusively in post-menopausal females, whereas no significant associations were observed in pre-menopausal females.

#### Overall psychosexual health

Among post-menopausal females, reporting poorer well-being (= higher scores in the wellPC), a stronger menopause-like endocrine profile, was associated with poorer overall psychosexual health. Whereas no such association was observed among those reporting better well-being. Thus, the endocrine milieu may be particularly relevant to psychosexual health in post-menopausal females with lower well-being. Conversely, better well-being may moderate the association between endocrine status and psychosexual health. This interpretation is consistent with evidence linking anxiety, depressive symptoms, and perceived stress to poorer sexual health throughout the reproductive life span (22,23,31,59), as well as evidence that resilience, better education and increased socioeconomic status is associated with better female sexual health (25,60,61). These relationships appear particularly pronounced during perimenopause, where greater depressive symptom severity has been linked to lower psychosexual health (25). Similarly, females with premature ovarian insufficiency who exhibit elevated depressive symptoms, anxiety, and stress, also demonstrate poorer psychosexual functioning alongside endocrine alterations such as elevated LH and reduced AMH levels (59).

#### Pain

In those post-menopausal females, with poorer (=higher wellPC scores) and mean scores in well-being (=mean wellPC scores), a stronger menopause-like endocrine profile was associated with less pain during sexual intercourse (= higher sexual pain scores). There was no association between the endocrine milieu and sexual pain in post-menopausal women reporting better well-being (= lower wellPC scores). No significant association was observed in pre-menopausal females. This is indicating that the endocrine milieu specifically impacts those post-menopausal females reporting low well-being.

A potential biological pathway linking the endocrine milieu to sexual pain involves vulvovaginal and genitourinary changes associated with menopause, including reduced lubrication, elasticity, and tissue integrity (48–51). Menopause-related endocrine changes have also been associated with greater sexual pain and vaginal discomfort (62). Also, genetic variation in hormone receptors (polymorphisms in the androgen receptor gene) have shown to be associated with sexual pain in midlife females (63). In addition, to a shift in hormone receptor density across different reproductive phases (52)this suggests that alterations in receptor expression or sensitivity may contribute to differences in the response to changes in the endocrine milieu. Our findings extend this literature by suggesting that the association between endocrine status and sexual pain may be particularly evident among post-menopausal females with poorer well-being.

The interaction between pain perception and well-being suggests that endocrine changes alone do not determine sexual health outcomes following menopause. Rather, well-being might additionally moderate the relationship between endocrine changes and sexual health. These results can partly explain the inconsistent literature regarding the role of reproductive hormones in female psychosexual health. While lower AMH concentrations have been associated with poorer psychosexual health during the reproductive years (64), other studies have reported no associations between individual hormones such as FSH or AMH and psychosexual health in peri-menopausal females (65). These previous studies have predominantly examined hormones in isolation rather than considering the endocrine milieu as an integrated system. Given that menopause represents a coordinated endocrine transition involving simultaneous changes across multiple hormones, multivariate endocrine profiles as assessed in the current study may provide a more biologically meaningful representation of menopausal status than individual hormone concentrations alone. Our findings further extend this perspective by demonstrating that the influence of endocrine changes on psychosexual health also depends on well-being.

### Multivariate endocrine and psychosocial profiles provide a more comprehensive framework for understanding psychosexual and psychophysiological health

Our findings suggest that multivariate representations of endocrine status and psychological well-being may provide a more comprehensive account of psychosexual and psychophysiological health than individual markers. Models incorporating endoPC × wellPC showed strong explanatory and predictive performance, suggesting that capturing the combined structure of endocrine and psychosocial domains may better reflect their relationship with psychosexual health than considering individual markers in isolation. This is consistent with evidence that integrating multiple biological domains can improve the characterisation of complex health outcomes (32–34).

This multivariate approach may be particularly relevant during menopausal transition, when reproductive aging involves coordinated changes across multiple endocrine pathways. Importantly, our findings do not suggest that individual biomarkers are uninformative, but rather that multivariate profiles may capture complementary information that is not apparent when markers are considered separately. More broadly, these findings support a biopsychosocial perspective in which psychophysiological and psychosexual health after menopause reflect interacting biological and psychological influences.

Future longitudinal and multimodal research should determine whether interventions that simultaneously address endocrine and psychosocial factors are particularly beneficial for females experiencing poorer psychological and sexual health during and after the menopausal transition.

### Limitations and future research

Although our findings indicate associations between endocrine profiles, well-being, psychophysiological symptoms, and psychosexual health, it cannot be determined whether endocrine changes contribute to changes in well-being and sexual functioning, whether psychological factors influence the experience of psychophysiological symptoms, or whether these relationships are reciprocal. Longitudinal studies with repeated assessments across the menopausal transition are needed to clarify these pathways.

Further, the well-being profile primarily captured negative or vulnerability-related indicators, including stress, depressive and anxiety symptoms, and reduced quality of life. Protective factors such as resilience, self-esteem, optimism, and adaptive emotion regulation were not comprehensively assessed. The absence of associations between well-being and some outcomes should therefore not be interpreted as evidence that psychological factors are unimportant. Future research should incorporate both vulnerability and protective dimensions of well-being to identify factors that may increase resilience to menopause-related changes. The present study did not comprehensively assess behavioural and interpersonal factors that may contribute to sexual health. In particular, sexual activity frequency, partnership status, and factors such as dyspareunia, lubricant use, pelvic floor function, and urinary symptoms may help explain individual differences in sexual functioning and the unexpected pattern observed for the FSFI pain domain. Their absence limits our ability to determine whether lower reported sexual pain reflects differences in the underlying experience of pain or differences in opportunities to experience and report sexual pain. Longitudinal studies incorporating repeated endocrine, psychological, behavioural, and relational assessments are therefore needed to disentangle these reciprocal pathways, identify resilience factors, and determine which females are most vulnerable or most resilient to menopause-related changes in sexual health and menopause associated symptoms.

Particular attention should be given to females with pre-existing mental health vulnerabilities, such as depression or anxiety, who may be especially susceptible to adverse changes in psychosexual and psychophysiological health and may therefore benefit most from early identification of endocrine profiles and targeted support. Future studies should also examine a broader spectrum of hormonal markers, as hormonal changes during the menopausal transition extend beyond gonadotropins and fertility-related markers to include substantial changes in oestradiol and progesterone. Characterising these broader endocrine profiles may be particularly important for understanding symptom trajectories and individual differences in response to hormone-based treatments, including the widely used menopausal hormone therapy. Such an approach may ultimately help identify females who are vulnerable to adverse changes in psychosexual and psychophysiological health and inform more personalised approaches to prevention and intervention.

## Conclusion

Taken together, our findings indicate that well-being, endocrine milieu, and psychosexual health are closely interconnected after menopause but not before menopause. Previous associations between individual reproductive hormones and well-being have been inconsistent (14,16), and our findings now add that female psychosexual health cannot be adequately understood by considering individual endocrine or subjective parameters in isolation. Rather, psychosexual health after menopause appears to emerge from the coordinated interplay between endocrine milieu and well-being. These findings highlight the value of a more comprehensive screening when approaching menopausal and psychosexual care by incorporating assessment of endocrine and well-being profiles. Integrating these assessments into clinical practice may enable earlier identification of females who may benefit from personalised treatment approaches supporting female health across the menopausal transition.

## List of abbreviations

FSH: follicle-stimulating hormone
LH: luteinizing hormone
AMH: anti-Müllerian hormone
MRS: menopausal rating scale
FSFI: female sexual functioning index
BDI-2: Becks depression index 2
WHO5: WHO five wellbeing index
PSS10: perceived stress scale 10
STAI: state and trait anxiety index
PCA: Principal component analysis
wellPC: well-being principal component
endoPC: endocrine principal component

## Declarations

### Ethics approval and consent to participate

Ethics Committee of the Medical Faculty of the University of Tuebingen gave ethical approval for this work. We confirm that all necessary participant consent has been obtained and the appropriate institutional forms have been archived, and that any participant/sample identifiers included were not known to anyone (e.g., hospital staff, patients, or participants themselves) outside the research group so cannot be used to identify individuals.

## Data availability

The datasets analysed during the current study are not publicly available due to participant privacy considerations but are available from the corresponding author on reasonable request.

## Code availability

The underlying code for this study is not publicly available but may be made available to qualified researchers on reasonable request from the corresponding author.

## Competing interest

Over the past five years, Inger Sundström Poromaa has received an unrestricted grant from Gedeon Richter for studies on hormonal contraceptives. In addition, she has received lecture fees from Gedeon Richter, Bayer A/G, Exeltis and Astellas. All other authors declare no financial or non-financial competing interests.

## Funding

This study was funded by the German Research Foundation (DFG, IRTG 2804). The funders played no role in study design, data collection, analysis and interpretation of data, or the writing of this manuscript.

## Data Availability

All data produced in the present study are available upon reasonable request to the authors

## Acknowledgements

We would like to especially thank everyone within the International Research Training Group IRTG2804 for their support.

## Author contributions

**AFD:** Data collection; Formal analysis; Data curation; Roles/Writing -original draft & review. **JS, SB, AD, AW, PB, AS:** Data collection; Roles/Writing – review & editing. **MH:** Supervision; Roles/Writing -review & editing**. ISP:** Conceptualization; Funding acquisition; Project administration; Supervision; Roles/Writing -review & editing. **BD**: Conceptualization; Funding acquisition; Project administration; Resources; Supervision; Roles/Writing -review & editing. **LK**: Conceptualization; Investigation; Data curation; Methodology; Project administration; Supervision; Validation; Visualization; Roles/Writing – original draft, review & editing.

## Declaration of AI use

During the preparation of this work the author(s) used ChatGPT (OpenAI, San Francisco, CA) to assist with language editing. After using this tool/service, the authors reviewed and edited the content as needed and take full responsibility for the content of the publication.

## Notes

### Author Declarations

Ethics Committee of the Medical Faculty of the University of Tuebingen gave ethical approval for this work.

### Summary of Updates

This version of the manuscript has been revised to update the following: refinement of wordning in title and text body.

